# The diagnostic and triage accuracy of digital and online symptom checker tools: a systematic review

**DOI:** 10.1101/2021.12.21.21268167

**Authors:** William Wallace, Calvin Chan, Swathikan Chidambaram, Lydia Hanna, Fahad Mujtaba Iqbal, Amish Acharya, Pasha Normahani, Hutan Ashrafian, Sheraz R Markar, Viknesh Sounderajah, Ara Darzi

## Abstract

**Objective:** To evaluate the accuracy of digital and online symptom checkers in providing diagnoses and appropriate triage advice.

**Design:** Systematic review.

**Data sources:** Medline and Web of Science were searched up to 15 February 2021.

**Eligibility criteria for study selection:** Prospective and retrospective cohort, vignette, or audit studies that utilised an online or application-based service designed to input symptoms and biodata in order to generate diagnoses, health advice and direct patients to appropriate services were included.

**Main outcome measures:** The primary outcomes were (1) the accuracy of symptom checkers for providing the correct diagnosis and (2) the accuracy of subsequent triage advice given.

**Data extraction and synthesis:** Data extraction and quality assessment (using the QUADAS-2 tool) were performed by two independent reviewers. Owing to heterogeneity of the studies, meta-analysis was not possible. A narrative synthesis of the included studies and pre-specified outcomes was completed.

**Results:** Of the 177 studies retrieved, nine cohort studies and one cross-sectional study met the inclusion criteria. Symptom checkers evaluated a variety of medical conditions including ophthalmological conditions, inflammatory arthritides and HIV. 50% of the studies recruited real patients, while the remainder used simulated cases. The diagnostic accuracy of the primary diagnosis was low (range: 19% to 36%) and varied between individual symptom checkers, despite consistent symptom data input. Triage accuracy (range: 48.8% to 90.1%) was typically higher than diagnostic accuracy. Of note, one study found that 78.6% of emergency ophthalmic cases were under-triaged.

**Conclusions:** The diagnostic and triage accuracy of symptom checkers are variable and of low accuracy. Given the increasing push towards population-wide digital health technology adoption, reliance upon symptom checkers in lieu of traditional assessment models, poses the potential for clinical risk. Further primary studies, utilising improved study reporting, core outcome sets and subgroup analyses, are warranted to demonstrate equitable and non-inferior performance of these technologies to that of current best practice.

**PROSPERO registration number:** CRD42021271022.

**SUMMARY BOXES:** *What is already known on this topic:* Chambers et al. (2019) have previously examined the evidence underpinning digital and online symptom checkers, including the accuracy of the diagnostic and triage information, for urgent health problems and found that diagnostic accuracy was generally low and varied depending on the symptom checker used. Given the increased reliance upon digital health technologies by health systems in light of the ongoing COVID-19 pandemic, in addition to the marked increase in availability of similarly themed digital health products since the last systematic review, a contemporary and comprehensive reassessment of this class of technologies to ascertain their diagnostic and triage accuracy is warranted.

*What this study adds:* Our systematic review demonstrates that the diagnostic accuracy of symptom checkers remains low and varies significantly depending on the pathology or symptom checker used. The findings of this systematic review suggests that this class of technologies, in their current state, poses significant risk for patient safety, particularly if utilised in isolation.

## INTRODUCTION

Digital and online symptom checkers (SCs) are application or software tools that enable patients to input their symptoms and biodata to produce a set of differential diagnoses and clinical triage advice. The diagnostic function of SCs is to provide a list of differential diagnoses, ranked by likelihood.[1] The triage function highlights to end-users the most appropriate course of action regarding their potential diagnosis, which typically includes seeking urgent care; contacting their general practitioner; or self-care. SCs have become an increasingly prominent feature of the modern healthcare landscape due to increasing access to internet connectivity and capacity to access personalised self-care advice. In 2020, 96% of UK households had internet access, of which over one-third of adults used the internet to self-diagnose health-related issues.[2,3] Governments have also incorporated SCs to alleviate the increasing burden that is placed upon both primary care services and emergency services, particularly in light of the COVID-19 pandemic.[4–6] It has been previously estimated that 12% of Emergency Department (ED) attendances would be more appropriately managed by other services.[7,8] Hence, SCs can reduce the financial and resource burden of the NHS, and redirect them towards truly in need. Public and private companies have advertised SCs to be a cost-effective solution by serving as a first port-of-call for patients and effectively signposting patients to the most appropriate healthcare service. When used appropriately, SCs can advise patients with serious conditions to seek urgent attention and conversely prevent those with problems best resolved through self-care from unnecessarily seeking medical attention.[1]

However, all of the aforementioned health, organisational and financial benefits of SCs are heavily dependent on the accuracy of diagnostic and triage advice provided. Over-triaging those with non-urgent ailments will exacerbate the unnecessary use of healthcare services. Conversely and more seriously, inaccuracies in diagnosing and triaging patients with life-threatening conditions could result in preventable morbidity and mortality.[1,9] In fact, SCs have previously received heavy media criticism for not correctly diagnosing cancer, cardiac conditions, and providing differing advice to patients with same symptomatology but different demographic characteristics.[10–12] These alleged reports raise concerns around the possibility that these systems may deliver unequitable clinical performance across differing gender and sociodemographic groups. In a previous systematic review, Chambers et al. (2019) assessed SCs on their safety and ability to correctly diagnose and distinguish between high and low acuity conditions.[13] The diagnostic accuracy was found to be variable between different platforms and was generally low. Given the rapid expansion in commercially available digital and online SCs, a more updated review is warranted to determine if this is still the case. Thus, this review aims to systematically evaluate the currently available literature regarding (1) the accuracy of digital and online SCs in providing diagnoses and appropriate triage advice as well as (2) the variation in recommendations provided by differing systems given homogenous clinical input data.

## METHODS

### Eligibility criteria

This systematic review was conducted according to the Preferred Reporting Items for Systematic Reviews and Meta-Analyses (PRISMA) guidelines and was registered in the PROSPERO registry (ID: CRD42021271022).[14] Prospective and retrospective cohort, vignette, or audit studies were included. Studies that utilised an online or application-based service designed to input symptoms and biodata (i.e., age and gender) in order to generate diagnoses, health advice and direct patients to appropriate services were included. All study populations, including patients, patient cases or simulated vignettes were included. Studies were included regardless of the condition(s) being assessed or the SC used. Included studies had to quantitatively evaluate the accuracy of the SC service. Excluded articles included descriptive studies, abstracts, commentaries, and study protocols. Only articles written in the English language were included.

### Search

Following PRISMA recommendations, an electronic database search was conducted using MEDLINE and Web of Science to include articles up to 15 February 2021 (search strategy detailed in supplementary text). Reference lists of the studies included in the review synthesis were examined for additional articles. Search results were then imported into Mendeley (RELX, UK) for duplicate removal and study selection. Screening of articles was performed independently by two investigators (W.W. and C.C.). Uncertainties were resolved through discussion with a third and fourth author (S.C and V.S).

### Data extraction and analysis

Key data were extracted and tabulated from the included studies, including details of study design, participants, interventions, and SCs used, comparators and reported study outcomes. Data extraction was performed independently by two investigators (W.W. and C.C.). The primary outcomes of this systematic review were (1) the accuracy of SCs for providing the correct diagnosis and (2) the accuracy of subsequent triage advice given (i.e., whether the acuity of the medical issue was correctly identified, and patients were signposted to appropriate services). The secondary outcome of assessing variation in recommendations within studies of consistent clinical input data can be calculated from these extracted outcomes. Due to heterogeneity of the included studies’ design, methodology and reported outcomes, a meta-analysis was not performed. A narrative synthesis of the included studies and pre-specified outcomes was instead carried out. Study bias was assessed using the Quality Assessment of Diagnostic Accuracy Studies (QUADAS-2) tool.[15] The risk of bias was assessed across the four domains by two investigators (W.W. and S.C.), any disagreements were discussed and resolved by a third author (V.S). The risk of bias of each domain was categorised as low, unclear, or high.

### Patient and public involvement

No patients were involved in the design or conduct of this study or writing and editing of the manuscript.

## RESULTS

### Search

The literature search yielded nine cohort studies and one cross-sectional study that met the inclusion criteria. Figure 1 presents the flow of studies through the screening process. An overview of the risk of bias assessment using QUADAS-2 can be found in Figure 2. All studies bar one had domains of “unclear” or “high” risk of bias or applicability concerns. Six studies had one or more domains at “high” risk of bias.[16–21]

**Figure 1.**
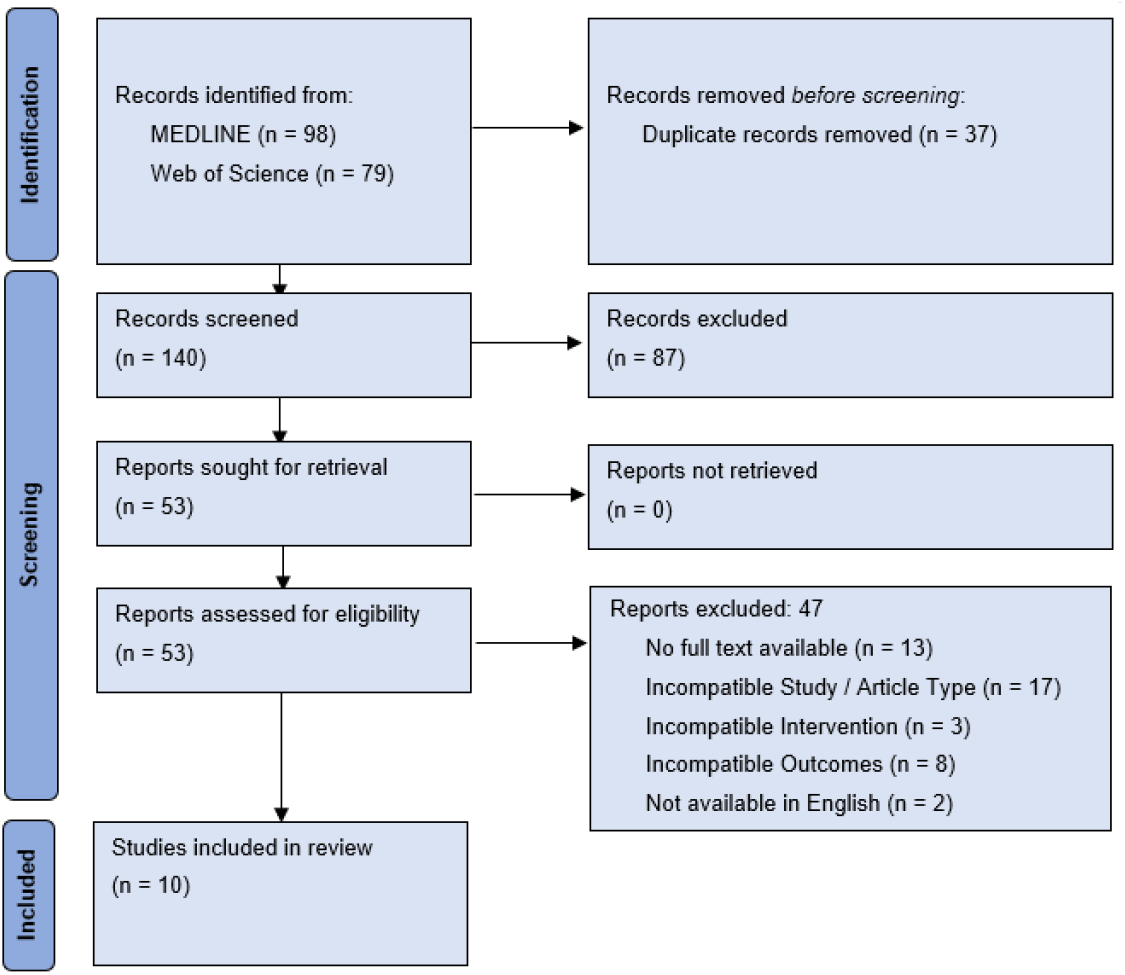
Preferred reporting items for Systematic Reviews and Meta-Analysis (PRISMA) flow diagram showing the process of study selection for this systematic review of symptom checker diagnostic and triage accuracy.

**Figure 2.**
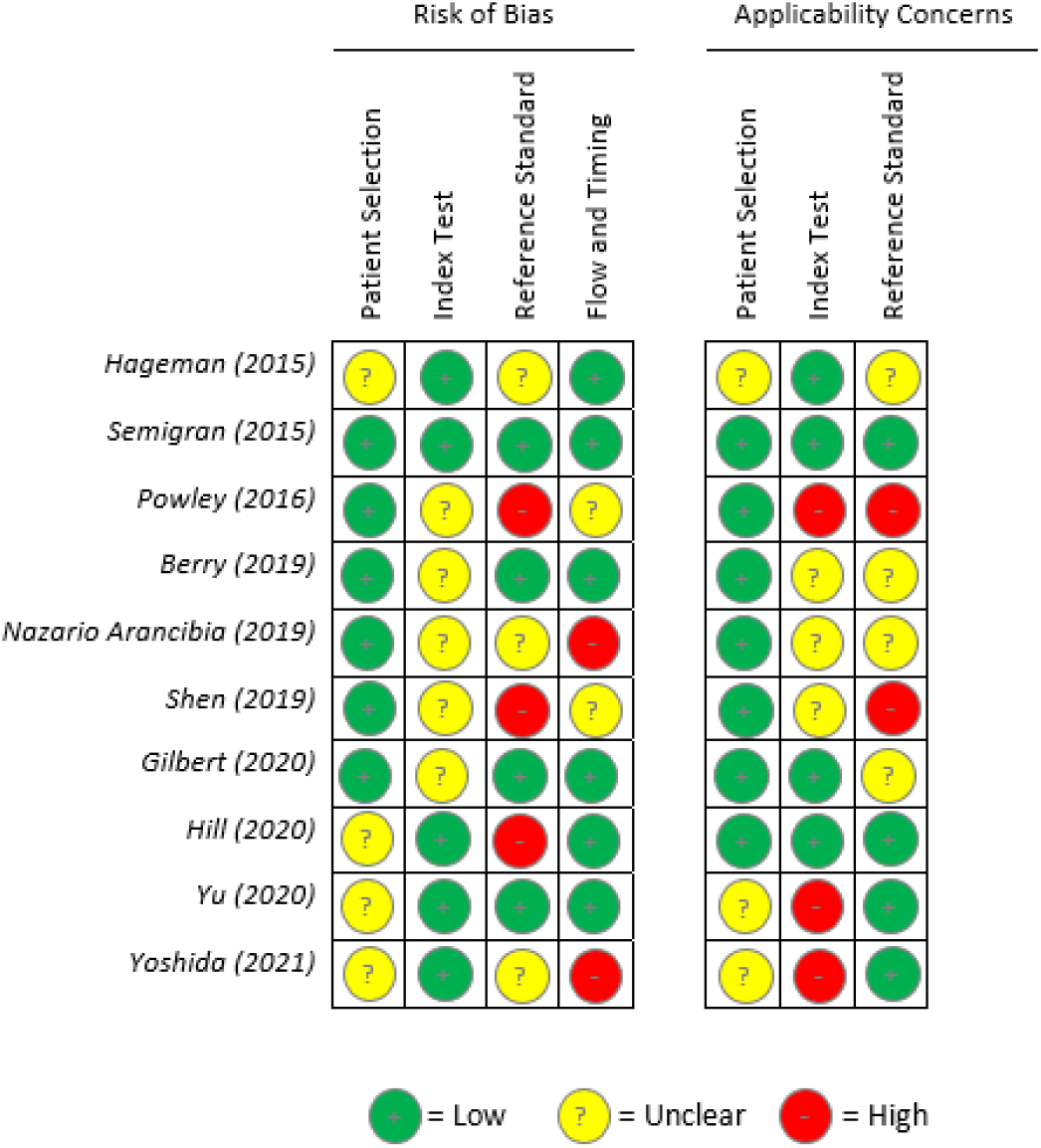
*Risk of bias summary using QUADAS-2 risk assessment tool*.[15] *Authors’ judgment regarding each domain of bias of each study synthesised on the accuracy of symptom checkers. Risk was categorised into one of three categories: Low risk (+), Unclear risk (?) and High risk (-)*.

### Characteristics of participants and interventions

Characteristics of included studies can be found in Table 1. Study population size ranged from 27 to 214 patients or vignettes.[17,21] Three studies were conducted in USA,[1,22,23] and one each in Australia,[19] Canada,[18] Hong Kong,[20] Spain,[17] and the United Kingdom.[16] The remaining two were multinational studies.[21,24]

**Table 1.**
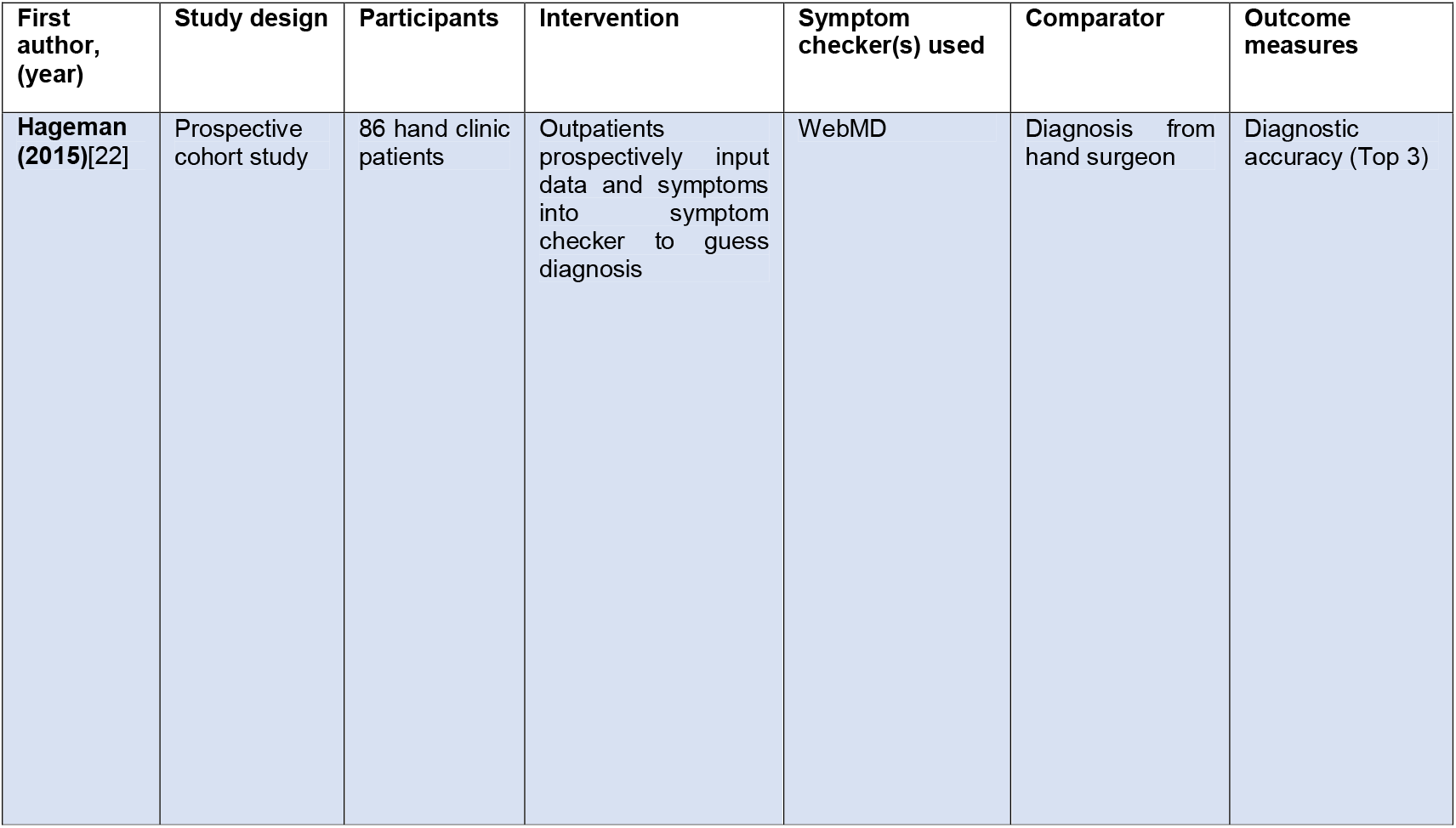
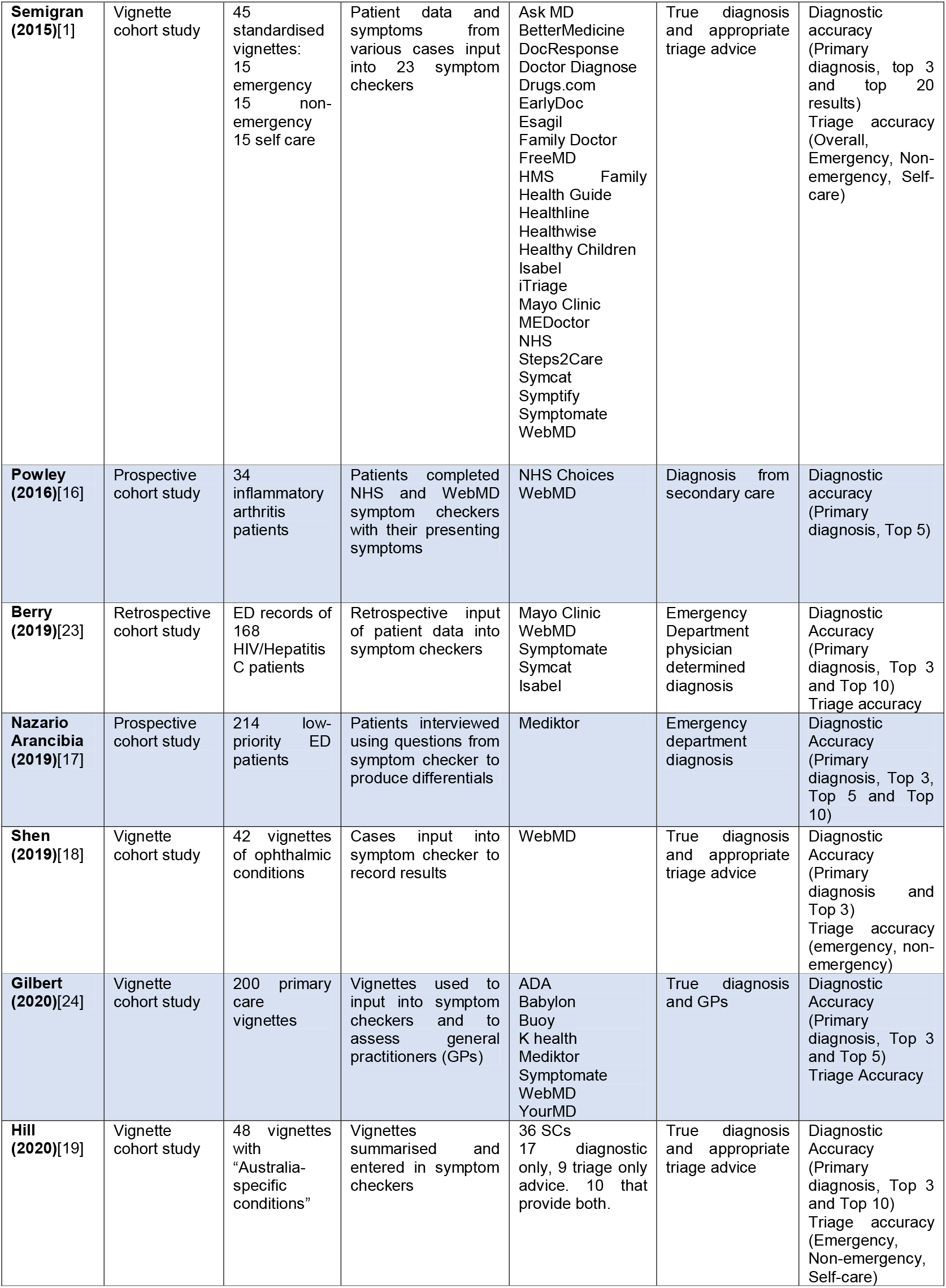
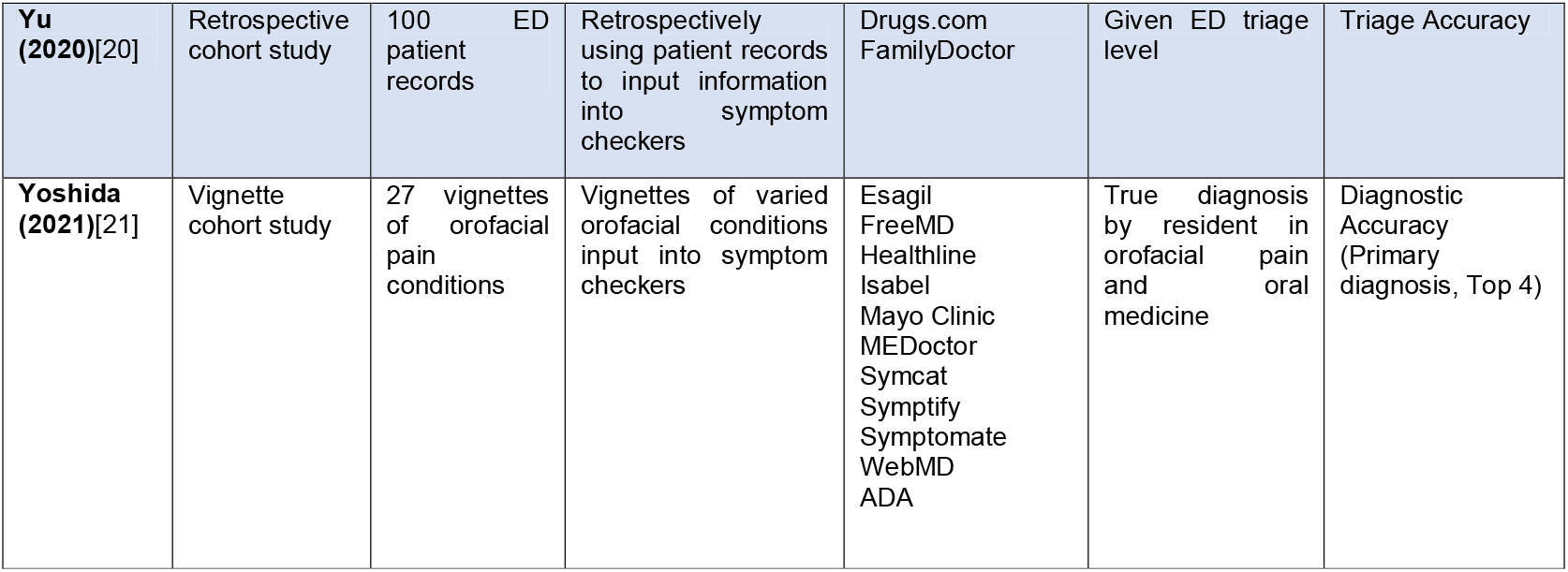
Characteristics of the ten studies included in the systematic review on the diagnostic and triage accuracy of symptom checkers.

Three studies involved prospective data collection (i.e., from outpatient clinics and ED waiting areas),[16,17,22] two studies involved retrospective analysis of ED clinical assessments,[20,23] and five studies used simulated patient vignettes.[1,18,19,21,24] The pathologies evaluated included hand conditions,[22] inflammatory arthritides,[16] infectious diseases (HIV and Hepatitis C),[23] ophthalmic conditions,[18] and orofacial conditions.[21] Four studies examined a wide range of general medical conditions pertinent to ED and general practice.[1,19,20,24]

A total of 48 different SCs were utilised in the included studies. Three studies only used one SC,[17,18,22] while two studies used more than 20 SCs.[1,19] Of note, the WebMD SC was assessed eight times and was the most commonly assessed SC in this review.[1,16,18,19,21–24]

### Accuracy of symptom checkers

#### Diagnostic accuracy

Nine studies evaluated SC diagnostic accuracy (Table 2). Overall primary diagnostic accuracy (i.e., listing the correct diagnosis first) was low in all studies, ranging from 19% to 38% (Figure 3).[16,17] Top three diagnostic accuracy, measured in seven studies, ranged from 33% to 58%.[17,22] Diagnostic accuracy for each specific SC can be found in Supplementary Table 1.

**Table 2.** Overall and range of average diagnostic and triage accuracy of symptom checkers in each study. Note. n/a – not applicable as only one symptom checker used; ns – not stated.

| First author (year) | No. of symptom checkers | Overall average diagnostic accuracy (%) | Range of average diagnostic accuracy (%) | Average triage accuracy (%) | Range of average triage accuracy (%) |
| --- | --- | --- | --- | --- | --- |
| Hageman (2015)[22] | 1 | 33 | n/a | ns | n/a |
| Semigran (2015)[1] | 23 | 34 | 5 – 50 | 57 | 33 – 78 |
| Powley (2016)[16] | 1 | 19 | n/a | ns | n/a |
| Berry (2019)[23] | 5 | ns | 3 – 16.4 | 48.8 | ns |
| Nazario Arancibia (2019)[17] | 1 | 37.9 | n/a | ns | n/a |
| Shen (2019)[18] | 1 | 26 | n/a | 66.7 | n/a |
| Gilbert (2020)[24] | 8 | 26.1 | 18 – 48 | 90.1 | 80 – 97.8 |
| Hill (2020)[19] | 36 | 36 | 12 – 61 | 49 | 17 – 61 |
| Yu (2020)[20] | 2 | ns | ns | 62 | 50 – 74 |
| Yoshida (2021)[21] | 11 | 21.7 | 0 – 38.5 | ns | ns |

**Figure 3.**
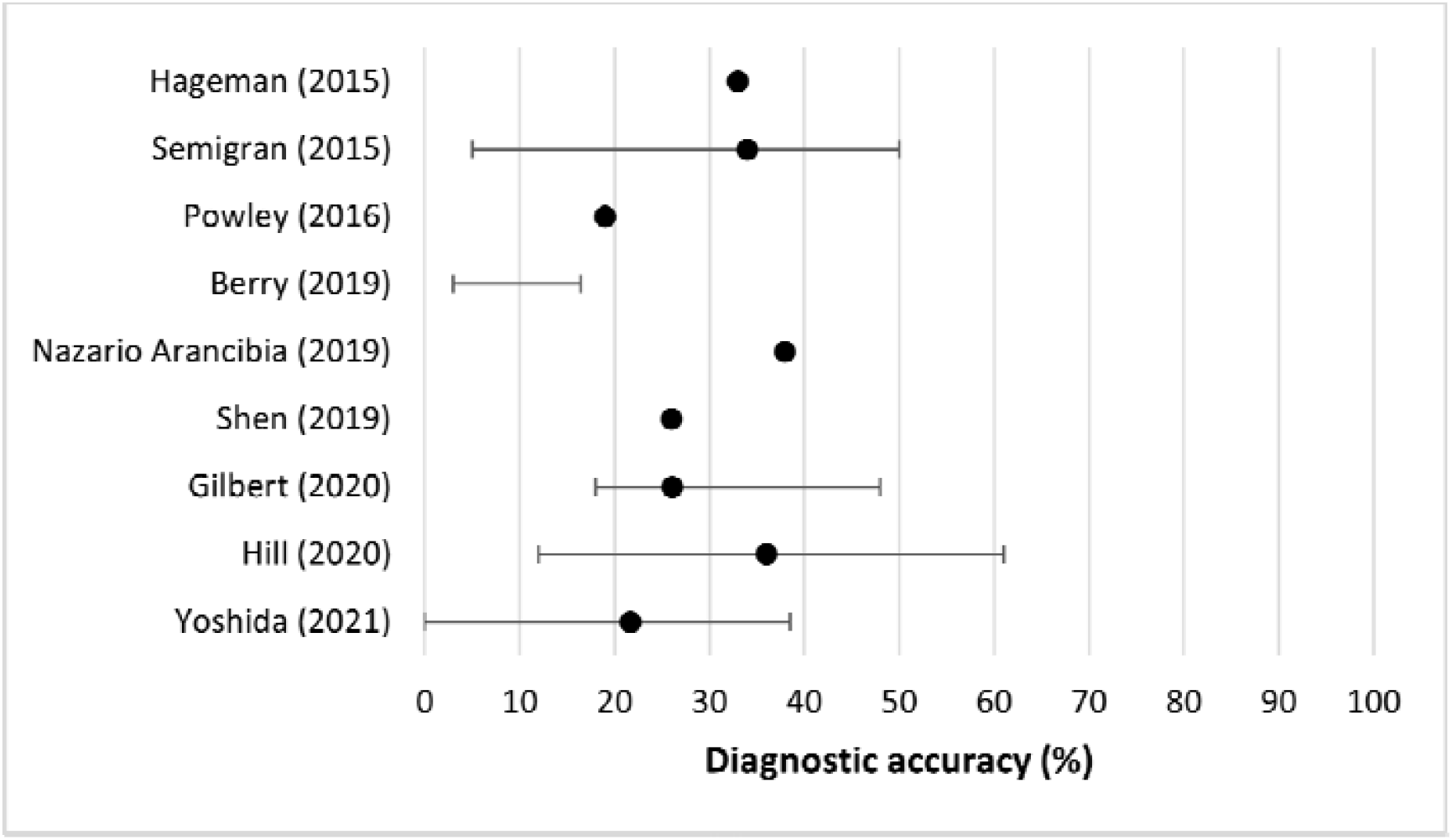
Mean primary diagnostic accuracy of symptom checkers in each study. Error bars signify the range of accuracy of different symptom checkers for the same patient/vignette population. Note. An overall accuracy value was not given in Berry (2019).

#### Triage accuracy

Six studies examined the accuracy of SCs in providing correct triage advice (Table 2). Overall triage accuracy tended to be higher than diagnostic accuracy, ranging from 49% to 90% (Figure 4).[19,23,24] Three studies stratified findings by emergency and non-emergency cases. Two studies reported that emergency cases were triaged more accurately (mean percentage [95% CI]) than non-urgent cases (80% [75-86] versus 55% [47-63], and 63% [52-71] versus 30% [11-39], respectively).[1,19] However, another study demonstrated that accuracy of triage advice for ophthalmic emergencies was significantly lower than non-urgent conditions (39% [14-64] versus 88% [73-100]).[18] Triage accuracy for each specific SC can be found in Supplementary Table 2.

**Figure 4.**
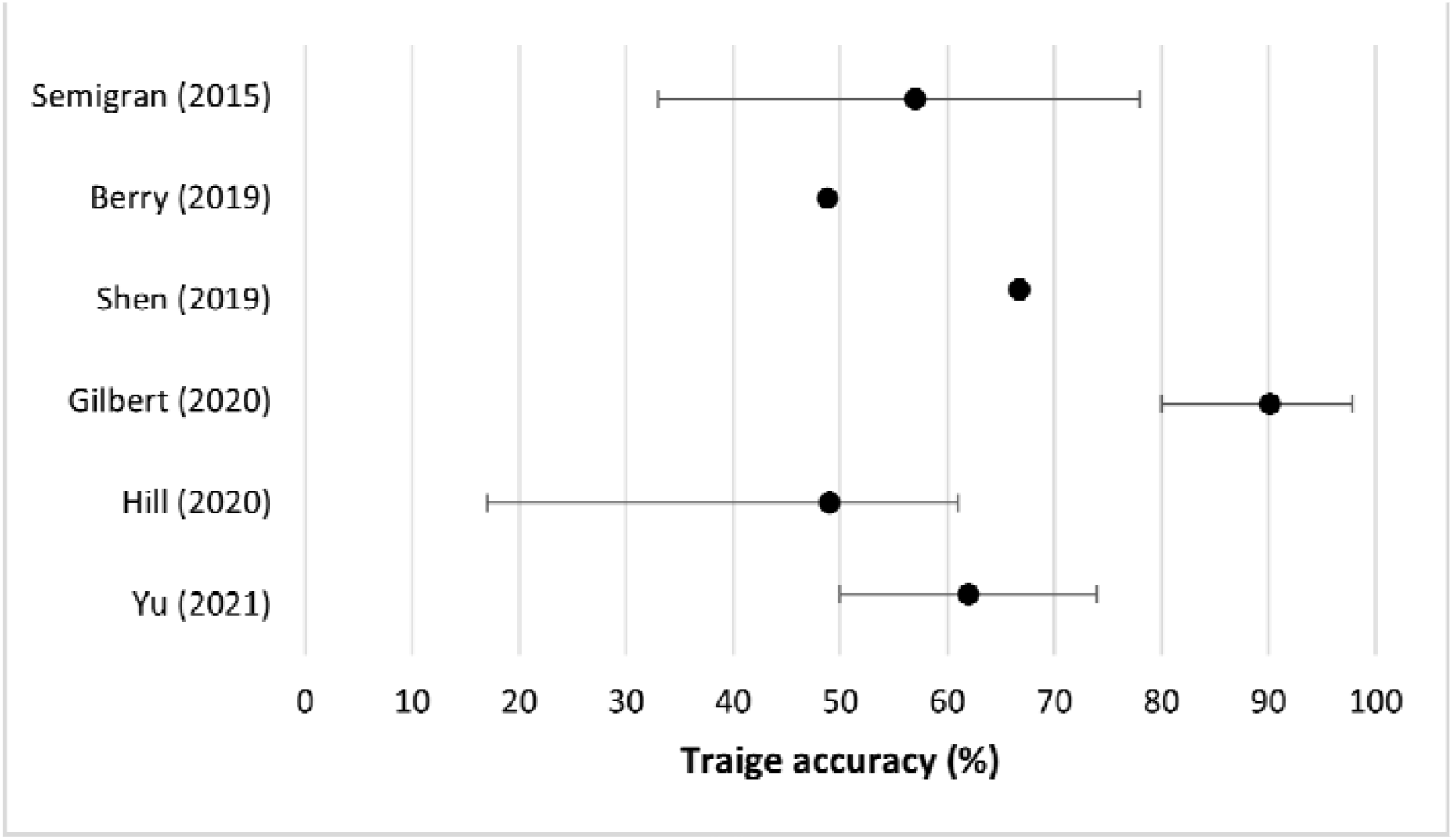
Mean accuracy of triage information given by symptom checkers in each study. Error bars signify the range of accuracy of different symptom checkers for the same patient/vignette population.

#### Variation in accuracy

All five studies that reported >1 SC demonstrated marked variability in diagnostic accuracy among different SCs for the same patient vignettes.[1,19,21,23,24] For a standardised set of general medical vignettes, mean primary diagnostic accuracy ranged from 5% to 50%.[1] Accuracy for diagnosing primary care conditions ranged from 18% to 48%.[24] Correct diagnosis of infectious diseases ranged from 3% to 16%.[23] Finally, primary diagnostic accuracy of different SCs assessing orofacial conditions ranged from 0% to 38.5%.[21] Similarly, the accuracy of triage advice given by individual SCs varied within a study. Semigran et al. and Hill et al. found a range of 33% to 78% and 17% to 61% respectively for general medical condtions.[1,19] Gilbert et al. reported a mean triage accuracy range of 80% to 97% for 200 primary care vignettes.[24] Variations in accuracy of providing diagnoses and triage advice was also apparent for specific SCs examining different conditions. The WebMD SC was most frequently used and was assessed in eight studies. The primary diagnostic accuracy of WebMD ranged from 3% to 36% across assessed conditions.[1,23] Shen et al. and Yoshida et al. found WebMD diagnostic accuracy to be 26% and 31% for ophthalmic and orofacial medicine cases respectively.[18,21]

## DISCUSSION

This systematic review evaluated the diagnostic and triage accuracy of symptom checkers for a variety of medical conditions using both simulated and real-life patient vignettes. Our review highlighted that both diagnostic and triage accuracy were generally low. Moreover, there is considerable variation in their performance despite consistencies in the input parameters. We also note that the diagnostic and triage accuracies of SCs, as well as the variation in performance, was greatly dependent on the acuity of the condition assessed. As a whole, these issues raise multiple concerns for the use of SCs as patient-facing tools, especially given their increasing role as triage services that direct patients towards appropriate treatment pathways.

Both SCs and telephone triage have been promoted as a means to reduce unnecessary GP and ED attendances. However, an inaccurate SC (one that does not suggest a correct set of diagnoses or provide safe triage advice), could expose patients to considerable preventable harm. When unsafe triage advice is paired with an incorrect set of differential diagnoses, this alignment of errors increases the likelihood for clinical harm of patients, not unlike the Swiss-Cheese model that is cited in aviation safety reports.[25] For example, Babylon, a NHS-backed SC, has been alleged to suggest that a breast lump may not necessarily represent cancer and it has also been reported to have misdiagnosed myocardial infarctions as panic attacks.[10,11] While there will be instances where probability-based clinical decision-making tools are incorrect, a safety-first approach needs to be employed for specific high-risk conditions with necessary adjustments for low-risk symptoms that may mask or mimic more life-threatening problems.

Variability in accuracy is a concerning recurrent theme in the included studies and indicates that patients are provided with heterogeneous advice, dependent on the SC used, and condition assessed, resulting in a spectrum of issues. Variability combined with poor diagnostic and triage accuracy presents a multidimensional system of potential patient harm. Although ‘undertriaging’ has clearly appreciable deleterious effects to patient wellbeing, it is worth noting that ‘overtriaging’ manifests in inappropriate health resource utilisation through unnecessary presentation to emergency services. Although this does not impact the health of the primary SC user, it does confer a knock-on opportunity cost that is shouldered by those are truly in need of emergency services and are left waiting for medical attention. Although the impact of variable triaging advice from SCs has yet to be robustly researched, the highly varied accuracy between SCs noted in this review suggests that there is considerable scope for discrepancies in quality and health outcomes. This raises further questions regarding the safety of SCs.

Many cite that the poor transparency and reporting of SC development and clinical validation limits the extent to which they can be reliably endorsed for population wide use between health systems. Minimal evidence is provided regarding the context, patient demographics and clinical information that is used to create SCs. This is reflected in the high risk of bias evident from the quality assessment of the included studies, with little elaboration of patient selection, comparator groups or index tests used. This can be improved by clearly stating what the intended use case and coverage of SCs will be. Coverage (i.e. what conditions and patient populations are accounted for by the software) must be explained, especially since SCs may not account for geographical or country-specific variations in disease prevalence, thus impacting applicability and potential utility. This could be further complemented with the open publication of algorithms, study protocols and datasets pertaining to SCs. Moreover, SCs currently do not display suitable explainability metrics, in which they highlight how they arrive to their recommendations. This would significantly increase the ability to effectively audit these devices as well as increase trust from both patients and healthcare professionals in the outputs they provide. Overall trust is also hindered by claims of several SCs to purported house ‘AI algorithms’ as part of their diagnostic process, despite not providing any convincing evidence as to how this is indeed the case.

### Limitations

Firstly, five of the studies used simulated patient vignettes, which are unlikely to represent the complexity of real patients.[26,27] Future work could include real-life patient vignettes to increase software exposure to the nuances of real-life patient populations. Obtaining a representative and balanced patient vignette set is especially pertinent for the training of SCs and development of AI algorithms. The lack of quality training data and the presence of dataset imbalances will likely directly influence the diagnostic model and will negatively impact diagnostic and triage accuracy.[28] Secondly, included retrospective designs introduce bias, reliant on the quality of documentation and knowledge of subsequent outcomes. Furthermore, this review did not capture all existing online SCs. The internet is abundant with SCs, and the majority do not have any validatory reports. With the expanding and evolving digital health market, newer and more improved SCs will be available. Future work should also evaluate the quality of these technologies and ensure they are fit for their purpose. Conversely, increased engagement with software developers to encourage reporting of diagnostic accuracy, including details of training and testing datasets, through standardised guidelines would allow direct comparison of efficacy and safety between different SCs. Lastly, there was a noticeable absence of middle and low economically developed countries, which are likely to exhibit different health seeking behaviours, digital literacy rates, and disease burden.

### Future direction and recommendations

Increased regulation of SCs via health technology assessments is essential given the wide impact of such potent technologies and has been advocated for previously.[9,29] In the UK, although the MHRA has recently outlined concerns regarding SCs, the long-awaited expansion of the current regulations for software-based medical products to adequately cover SCs has not yet been realised.[30] Increased regulatory scrutiny is greatly needed in the near future given the rapidly-progressing nature of this field. Robust clinical validation and testing with is warranted to improve current software trustworthiness and reliability. Digital health studies that form the basis for SCs need to be carried out with greater methodological rigour and transparency. This can be achieved by incorporating core outcome sets; real-world patient data encompassing greater demographic spread, particularly ethnicity which is not captured in the shortlisted studies despite often being a key risk factor in many pathologies; and more refined comparator groups that include biochemical, clinical, and radiological diagnosis when feasible. While SCs fulfil the need for telemedicine, further work should also evaluate whether SCs truly are better than more traditional telephone triage lines, especially in terms of cost-effectiveness, as this service also provides ‘socially distanced’ and personalised health information. More importantly, there is an unmet need for educating patients in using these tools and appreciating their limitations. While the variation in digital health literacy has previously been established, more effort is required to address and correct its socio-economic drivers.

## CONCLUSION

In our review, SC diagnostic and triage accuracy varied substantially and was generally low. Variation exists between different SCs and the conditions being assessed; this raises safety and regulatory concerns. Given the increasing trend of telemedicine use and, even the endorsement of certain applications by the NHS, further work should seek to introduce regulation and establish datasets to support their development and improve patient safety.

## Data Availability

All data produced in the present work are contained in the manuscript

## CONTRIBUTORS

William Wallace, Calvin Chan and Swathikan Chidambaram contributed to the data collection and analysis. Lydia Hanna, Amish Acharya, Fahad Iqbal, Pasha Normahani and contributed to manuscript writing. Hutan Ashrafian, Sheraz Markar, Viknesh Sounderajah and Ara Darzi contributed to the critical revision of the manuscript as well as initial study conception. Viknesh Sounderajah is the guarantor of the study. The corresponding author attests that all listed authors meet authorship criteria and that no others meeting the criteria have been omitted.

## LICENSE FOR PUBLICATION

The Corresponding Author has the right to grant on behalf of all authors and does grant on behalf of all authors, a worldwide licence to the Publishers and its licensees in perpetuity, in all forms, formats and media (whether known now or created in the future), to i) publish, reproduce, distribute, display and store the Contribution, ii) translate the Contribution into other languages, create adaptations, reprints, include within collections and create summaries, extracts and/or, abstracts of the Contribution, iii) create any other derivative work(s) based on the Contribution, iv) to exploit all subsidiary rights in the Contribution, v) the inclusion of electronic links from the Contribution to third party material where-ever it may be located; and, vi) licence any third party to do any or all of the above.

## ETHICAL APPROVAL

Not required.

## TRANSPARANCY STATEMENT

The lead author (the manuscript’s guarantor) affirms that the manuscript is an honest, accurate, and transparent account of the study being reported; that no important aspects of the study have been omitted; and that any discrepancies from the study as planned (and, if relevant, registered) have been explained.

## DATA SHARING

The search strategy is available in the supplementary material; any additional data are available on request.

---

PRISMA 2020 Checklist
| Section and Topic | Item # | Checklist item | Page where item is reported |
| --- | --- | --- | --- |
| **TITLE** |  |  |  |
| Title | 1 | Identify the report as a systematic review. | 1 |
| **ABSTRACT** |  |  |  |
| Abstract | 2 | See the PRISMA 2020 for Abstracts checklist. | 3 |
| **INTRODUCTION** |  |  |  |
| Rationale | 3 | Describe the rationale for the review in the context of existing knowledge. | 5 |
| Objectives | 4 | Provide an explicit statement of the objective(s) or question(s) the review addresses. | 5 |
| **METHODS** |  |  |  |
| Eligibility criteria | 5 | Specify the inclusion and exclusion criteria for the review and how studies were grouped for the syntheses. | 6 |
| Information sources | 6 | Specify all databases, registers, websites, organisations, reference lists and other sources searched or consulted to identify studies. Specify the date when each source was last searched or consulted. | 6 |
| Search strategy | 7 | Present the full search strategies for all databases, registers and websites, including any filters and limits used. | Suppl text |
| Selection process | 8 | Specify the methods used to decide whether a study met the inclusion criteria of the review, including how many reviewers screened each record and each report retrieved, whether they worked independently, and if applicable, details of automation tools used in the process. | 6 |
| Data collection process | 9 | Specify the methods used to collect data from reports, including how many reviewers collected data from each report, whether they worked independently, any processes for obtaining or confirming data from study investigators, and if applicable, details of automation tools used in the process. | 6 |
| Data items | 10a | List and define all outcomes for which data were sought. Specify whether all results that were compatible with each outcome domain in each study were sought (e.g. for all measures, time points, analyses), and if not, the methods used to decide which results to collect. | 6 |
|  | 10b | List and define all other variables for which data were sought (e.g. participant and intervention characteristics, funding sources). Describe any assumptions made about any missing or unclear information. | 6 |
| Study risk of bias assessment | 11 | Specify the methods used to assess risk of bias in the included studies, including details of the tool(s) used, how many reviewers assessed each study and whether they worked independently, and if applicable, details of automation tools used in the process. | 6 |
| Effect measures | 12 | Specify for each outcome the effect measure(s) (e.g. risk ratio, mean difference) used in the synthesis or presentation of results. | 6 |
| Synthesis methods | 13a | Describe the processes used to decide which studies were eligible for each synthesis (e.g. tabulating the study intervention characteristics and comparing against the planned groups for each synthesis (item #5)). | 6 |
|  | 13b | Describe any methods required to prepare the data for presentation or synthesis, such as handling of missing summary statistics, or data conversions. | 6 |
|  | 13c | Describe any methods used to tabulate or visually display results of individual studies and syntheses. | 6 |
|  | 13d | Describe any methods used to synthesize results and provide a rationale for the choice(s). If meta-analysis was performed, describe the model(s), method(s) to identify the presence and extent of statistical heterogeneity, and software package(s) used. | 6 |
|  | 13e | Describe any methods used to explore possible causes of heterogeneity among study results (e.g. subgroup analysis, meta-regression). | n/a |
|  | 13f | Describe any sensitivity analyses conducted to assess robustness of the synthesized results. | n/a |
| Reporting bias assessment | 14 | Describe any methods used to assess risk of bias due to missing results in a synthesis (arising from reporting biases). | 6 |
| Certainty | 15 | Describe any methods used to assess certainty (or confidence) in the body of evidence for an outcome. | 6 |

PRISMA 2020 Checklist
| Section and Topic | Item # | Checklist item | Page where item is reported |
| --- | --- | --- | --- |
| assessment |  |  |  |
| **RESULTS** |  |  |  |
| Study selection | 16a | Describe the results of the search and selection process, from the number of records identified in the search to the number of studies included in the review, ideally using a flow diagram. | 7 |
|  | 16b | Cite studies that might appear to meet the inclusion criteria, but which were excluded, and explain why they were excluded. | 7 |
| Study characteristics | 17 | Cite each included study and present its characteristics. | Table 1 |
| Risk of bias in studies | 18 | Present assessments of risk of bias for each included study. | Figure 2 |
| Results of individual studies | 19 | For all outcomes, present, for each study: (a) summary statistics for each group (where appropriate) and (b) an effect estimate and its precision (e.g. confidence/credible interval), ideally using structured tables or plots. | Table 2 |
| Results of syntheses | 20a | For each synthesis, briefly summarise the characteristics and risk of bias among contributing studies. | n/a |
|  | 20b | Present results of all statistical syntheses conducted. If meta-analysis was done, present for each the summary estimate and its precision (e.g. confidence/credible interval) and measures of statistical heterogeneity. If comparing groups, describe the direction of the effect. | n/a |
|  | 20c | Present results of all investigations of possible causes of heterogeneity among study results. | 7 |
|  | 20d | Present results of all sensitivity analyses conducted to assess the robustness of the synthesized results. | n/a |
| Reporting biases | 21 | Present assessments of risk of bias due to missing results (arising from reporting biases) for each synthesis assessed. | 7 |
| Certainty of evidence | 22 | Present assessments of certainty (or confidence) in the body of evidence for each outcome assessed. | 7 |
| **DISCUSSION** |  |  |  |
| Discussion | 23a | Provide a general interpretation of the results in the context of other evidence. | 9 |
|  | 23b | Discuss any limitations of the evidence included in the review. | 10 |
|  | 23c | Discuss any limitations of the review processes used. | 10 |
|  | 23d | Discuss implications of the results for practice, policy, and future research. | 10 |
| **OTHER INFORMATION** |  |  |  |
| Registration and protocol | 24a | Provide registration information for the review, including register name and registration number, or state that the review was not registered. | 6 |
|  | 24b | Indicate where the review protocol can be accessed, or state that a protocol was not prepared. | 6 |
|  | 24c | Describe and explain any amendments to information provided at registration or in the protocol. | n/a |
| Support | 25 | Describe sources of financial or non-financial support for the review, and the role of the funders or sponsors in the review. | 12 |
| Competing interests | 26 | Declare any competing interests of review authors. | 12 |
| Availability of data, code and other materials | 27 | Report which of the following are publicly available and where they can be found: template data collection forms; data extracted from included studies; data used for all analyses; analytic code; any other materials used in the review. | 12 |

