## Supplementary material for "The diagnostic and triage accuracy of digital and online symptom checker tools: a systematic review": Tables

Table 1. Diagnostic accuracy for all studies. Overall accuracy along with individual symptom checker accuracy has been included for all measures of accuracy used in each study. Values depicted are means with additional representations of standard deviation (±), range ((…)) and 95% confidence intervals ([…]).

| First Author (Year) | Symptom Checker | | Mean Diagnostic Accuracy (%) for different accuracy measures | | | | | |
| --- | --- | --- | --- | --- | --- | --- | --- | --- |
|  |  |  | Primary Diagnosis | Top 3 | Top 4 | Top 5 | Top 10 | Top 20 |
| Hageman (2015) | Overall (WebMD) | |  | 33.0 |  |  |  |  |
| Semigran (2015) | Overall | | 34.0  [31-37] | 51.0  [47-54] |  |  |  | 58.0  [55-62] |
|  | Individual | Ask MD | 43 | 68 |  |  |  | 75 |
|  |  | BetterMedicine | 24 | 29 |  |  |  | 38 |
|  |  | DocResponse | 50 | 67 |  |  |  | 72 |
|  |  | Doctor Diagnose | 41 | 44 |  |  |  | 46 |
|  |  | Drugs.com | 40 | 47 |  |  |  | 58 |
|  |  | EarlyDoc | 32 | 33 |  |  |  | 33 |
|  |  | Esagil | 20 | 34 |  |  |  | 50 |
|  |  | Family Doctor | 47 | 56 |  |  |  | 56 |
|  |  | FreeMD | 36 | 45 |  |  |  | 48 |
|  |  | HMS FHG* | 34 | 52 |  |  |  | 55 |
|  |  | Healthline | 38 | 53 |  |  |  | 58 |
|  |  | Isabel | 44 | 69 |  |  |  | 84 |
|  |  | iTriage | 36 | 64 |  |  |  | 77 |
|  |  | Mayo Clinic | 17 | 59 |  |  |  | 76 |
|  |  | MEDoctor | 5 | 43 |  |  |  | 43 |
|  |  | Symcat | 40 | 71 |  |  |  | 76 |
|  |  | Symptify | 29 | 36 |  |  |  | 44 |
|  |  | Symptomate | 31 | 34 |  |  |  | 34 |
|  |  | WebMD | 36 | 51 |  |  |  | 62 |
| Powley (2016) | Overall (WebMD) | | 19.0 |  |  | 71.0 |  |  |
| Berry (2019) | Overall | | <20.0 | <35.0 |  |  | <40.0 |  |
|  | Individual  (Both HIV and Hep C) | Symptomate | 7.1 | 7.7 |  |  | 8.9 |  |
|  |  | Symcat | 11.3 | 26.9 |  |  | 32.1 |  |
|  |  | Isabel | 7.1 | 16.1 |  |  | 25.6 |  |
|  |  | Mayo Clinic | 6.0 | 13.1 |  |  | 22.0 |  |
|  |  | WebMD | 7.1 | 17.3 |  |  | 28.0 |  |
|  | Individual  (HIV alone) | Symptomate | 5.6 | 5.6 |  |  | 5.6 |  |
|  |  | Symcat | 6.7 | 20.0 |  |  | 26.7 |  |
|  |  | Isabel | 5.6 | 17.8 |  |  | 28.9 |  |
|  |  | Mayo Clinic | 4.4 | 10.0 |  |  | 22.2 |  |
|  |  | WebMD | 7.8 | 16.7 |  |  | 26.7 |  |
|  | Individual  (Hep C alone) | Symptomate | 7.5 | 9.0 |  |  | 11.9 |  |
|  |  | Symcat | 16.4 | 32.8 |  |  | 37.3 |  |
|  |  | Isabel | 7.5 | 13.4 |  |  | 20.9 |  |
|  |  | Mayo Clinic | 6.0 | 14.9 |  |  | 20.9 |  |
|  |  | WebMD | 3.0 | 14.9 |  |  | 23.9 |  |
| Nazario Arancibia (2019) | Overall (Mediktor) | | 37.9 | 58.0 |  | 65.4 | 76.5 |  |
| Shen (2019) | Overall (WebMD) | | 26.0  [12-40] | 38.0  [25-56] |  |  |  |  |
| Gilbert (2020) | Overall |  | 26.1±8.9 | 38.0±13.5 |  | 40.8±15.2 |  |  |
|  | Individual | Ada | 48.0 | 70.5 |  | 77.0 |  |  |
|  |  | Babylon | 22.0 | 32.0 |  | 32.0 |  |  |
|  |  | Buoy | 24.0 | 43.0 |  | 43.0 |  |  |
|  |  | K health | 25.0 | 36.0 |  | 39.0 |  |  |
|  |  | Mediktor | 24.0 | 36.0 |  | 37.0 |  |  |
|  |  | Symptomate | 18.0 | 27.5 |  | 30.0 |  |  |
|  |  | WebMD | 20.0 | 35.5 |  | 44.0 |  |  |
|  |  | YourMD | 21.0 | 23.5 |  | 24.0 |  |  |
| Hill (2020) | Overall | | 36.0  [31-42] | 52.0  [47-59] |  | 58.0  [53-65] |  |  |
|  | Individual | AARP Health Tools | 31 | 38 |  | 40 |  |  |
|  |  | American Postal Workers Union Health Plan | 25 | 35 |  | 40 |  |  |
|  |  | Buoy Health | 35 | 50 |  | 50 |  |  |
|  |  | Drugs.com | 37 | 56 |  | 56 |  |  |
|  |  | Everyday Health | 52 | 71 |  | 71 |  |  |
|  |  | Family Doctor | 40 | 46 |  | 46 |  |  |
|  |  | Healthline | 27 | 46 |  | 60 |  |  |
|  |  | Health Status | 37 | 59 |  | 73 |  |  |
|  |  | Isabel | 33 | 63 |  | 77 |  |  |
|  |  | Mayo Clinic | 38 | 60 |  | 67 |  |  |
|  |  | MedicineNet | 27 | 54 |  | 67 |  |  |
|  |  | Patient.info | 44 | 56 |  | 63 |  |  |
|  |  | Right Diagnosis | 14 | 36 |  | 41 |  |  |
|  |  | RxList | 31 | 58 |  | 71 |  |  |
|  |  | Symcat | 37 | 48 |  | 65 |  |  |
|  |  | Symptomate | 61 | 77 |  | 81 |  |  |
|  |  | WebMD | 53 | 70 |  | 77 |  |  |
|  |  | What’s My Diagnosis | 19 | 23 |  | 30 |  |  |
|  |  | Doctor Diagnose (Google) | 33 | 44 |  | 44 |  |  |
|  |  | Drugs.com (Google) | 44 | 58 |  | 58 |  |  |
|  |  | ePain Assist (Apple) | 14 | 28 |  | 40 |  |  |
|  |  | ePain Assist (Google) | 12 | 23 |  | 33 |  |  |
|  |  | Symptify (Google) | 24 | 30 |  | 30 |  |  |
|  |  | Symptomate (Apple) | 61 | 77 |  | 81 |  |  |
|  |  | Symptomate (Google) | 61 | 77 |  | 81 |  |  |
|  |  | WebMD (Apple) | 51 | 68 |  | 74 |  |  |
|  |  | WebMD (Google) | 53 | 70 |  | 77 |  |  |
| Yoshida (2021) | Overall |  | 21.7  (0-38.5) |  | 25.9  (3.9-38.5) |  |  |  |
|  | Individual | Ada | 26.9 |  | 34.6 |  |  |  |
|  |  | Esagil | 0 |  | 3.9 |  |  |  |
|  |  | FreeMD | 23.1 |  | 26.9 |  |  |  |
|  |  | Healthline | 38.5 |  | 42.3 |  |  |  |
|  |  | Isabel | 23.1 |  | 30.8 |  |  |  |
|  |  | Mayo Clinic | 7.7 |  | 7.7 |  |  |  |
|  |  | MEDoctor | 19.2 |  | 19.2 |  |  |  |
|  |  | Symcat | 30.8 |  | 38.5 |  |  |  |
|  |  | Symptify | 26.9 |  | 30.8 |  |  |  |
|  |  | Symptomate | 11.5 |  | 11.5 |  |  |  |
|  |  | WebMD | 30.8 |  | 38.5 |  |  |  |

Table 2. Triage accuracy of studies included in this review. Overall accuracy along with individual symptom checker accuracy has been included for all measures of accuracy used in each study. Values depicted are means with additional representations of standard deviation (±), range ((…)) and 95% confidence intervals ([…]).

| First Author (Year) | Symptom Checker | | Mean Triage Accuracy (%) | | | |
| --- | --- | --- | --- | --- | --- | --- |
|  |  |  | Overall Triage | Emergency Cases | Non-Emergency Cases | Self-care |
| Semigran (2015) | Overall | | 57.0 [52-61] | 80.0 [75-86] | 55.0 [47-63] | 33.0 [26-40] |
|  | Individual | Doctor Diagnose | 63 | 80 | 67 | 0 |
|  |  | Drugs.com | 60 | 57 | 60 | 62 |
|  |  | EarlyDoc | 53 | 67 | 60 | 33 |
|  |  | Family Doctor | 54 | 50 | 50 | 60 |
|  |  | FreeMD | 59 | 67 | 87 | 21 |
|  |  | HMS FHG | 78 | 93 | 85 | 62 |
|  |  | Healthwise | 43 | 100 | 7 | 21 |
|  |  | Healthy Children | 73 | 100 | 100 | 43 |
|  |  | Isabel | 51 | 100 | 53 | 0 |
|  |  | iTriage | 33 | 100 | 0 | 0 |
|  |  | NHS | 52 | 87 | 20 | 50 |
|  |  | Steps2Care | 71 | 86 | 71 | 57 |
|  |  | Symcat | 44 | 53 | 80 | 0 |
|  |  | Symptify | 70 | 92 | 71 | 50 |
|  |  | Symptomate | 64 | 76 | 67 | 0 |
| Berry (2019) | Overall | | 48.8 |  |  |  |
|  | Both HIV and Hepatitis C | | 45.5 |  |  |  |
|  | HIV alone | | 35.6 |  |  |  |
|  | Hepatitis C alone | | 59.7 |  |  |  |
| Shen (2019) | Overall (WebMD) | | 66.7 | 39.0 [14-64] | 88.0 [73-100] |  |
| Gilbert (2020) | Overall | | 90.1±7.4 |  |  |  |
|  | Individual | Ada | 97.0 |  |  |  |
|  |  | Babylon | 95.1 |  |  |  |
|  |  | Buoy | 80.0 |  |  |  |
|  |  | K Health | 81.3 |  |  |  |
|  |  | Mediktor | 87.3 |  |  |  |
|  |  | Symptomate | 97.8 |  |  |  |
|  |  | Your.MD | 92.6 |  |  |  |
| Hill (2020) | Overall | | 49.0  [44-54] | 63.0 [52-71] | 30.0 [11-39] | 40.0 [26-49] |
|  | Individual | Alberta Healthwise | 56 | 62 | 18 | 70 |
|  |  | Children’s Hospital of Wisconsin | 47 | 50 | 0 | 57 |
|  |  | Drugs.com | 51 | 54 | 75 | 25 |
|  |  | Everyday Health | 55 | 82 | 17 | 0 |
|  |  | Family Doctor | 31 | 38 | 45 | 10 |
|  |  | Healthdirect | 61 | 75 | 36 | 60 |
|  |  | Health Link BC | 54 | 62 | 18 | 60 |
|  |  | Healthy Children | 47 | 50 | 0 | 57 |
|  |  | Isabel | 48 | 100 | 27 | 0 |
|  |  | John Hopkins All Children’s Hospital | 47 | 50 | 0 | 57 |
|  |  | Michigan Medicine | 58 | 64 | 18 | 70 |
|  |  | St Luke’s Online | 47 | 50 | 0 | 57 |
|  |  | Symcat | 40 | 54 | 100 | 0 |
|  |  | Symptomate | 52 | 82 | 0 | 33 |
|  |  | Doctor Diagnose (Google) | 17 | 8 | 0 | 0 |
|  |  | Drugs.com (Google) | 53 | 54 | 88 | 25 |
|  |  | Healthdirect (Apple) | 61 | 75 | 36 | 60 |
|  |  | Symptomate (Apple) | 52 | 82 | 0 | 33 |
|  |  | Symptomate (Google) | 52 | 82 | 0 | 33 |
| Yu (2020) | Individual | Drugs.com | 74 |  |  |  |
|  |  | FamilyDoctor | 50 |  |  |  |

**Search terms:**

1. app*1 or application* or automated or artificial intelligence or AI or chatbot or computer or computer-based or computer-assisted or online or digital or web or website or mobile or phone or internet
2. diagnos?s or diagnos* or triage*
3. symptom* checker* or symptom assessment app or babylon or webmd or buoy or mediktor or symptomate or yourmd
4. 1 and 2 and 3
